# Time in blood glucose range 70 to 180 mg/dL and survival rate in critically ill patients: A retrospective observational study

**DOI:** 10.1101/2020.08.31.20184994

**Authors:** Hiromu Naraba, Tadahiro Goto, Toru Shirakawa, Tomohiro Sonoo, Naoki Kanda, Hidehiko Nakano, Yuji Takahashi, Hideki Hashimoto, Kensuke Nakamura

## Abstract

**Objective:** Time in targeted blood glucose range (TIR) 70-140 mg/dL has been associated with an increased risk of mortality in critically ill patients. Nevertheless, it remains unclear whether TIR is associated with 28-day mortality in critically ill patients under glycemic control with a less tight target glucose range of 70-180 mg/dL. We aimed to assess whether TIR 70-180 mg/dL was associated with 28-day mortality and to identify the optimal TIR.

**Design:** A retrospective observational study.

**Setting:** Data from a tertiary care centre in Japan, from 1 January 2016 through 31 October 2019.

**Participants:** 1,230 adult patients admitted to the intensive care unit for more than three days.

**Outcome measure:** The primary outcome was 28-day mortality.

**Results:** Of 1,230 patients, patients with HbA1c ≥6.5% had a higher 28-day mortality than those with <6.5% (32.0% vs. 22.7%; p=0.003). In the multivariate logistic regression, TIR <80% was associated with an increased risk of 28-day mortality in patients with HbA1c <6.5% with an adjusted odds ratio (OR) of 1.88 (95% confidence interval [CI]: 1.36-2.61). When using 10% incremental TIR as a categorical variable, lower TIR was associated with worse 28-day mortality compared to TIR ≥90% in patients with HbA1c <6.5% (e.g., adjusted OR of TIR <60%, 3.62 [95%CI 2.36-5.53]). Similar associations were found in the analyses using the COX proportional hazards model. In addition, sensitivity analyses using TIR of the first three days showed that the overall associations were consistent with primary analyses.

**Conclusions:** Our study demonstrated that lower TIR 70-180 mg/dL was associated with higher 28-day mortality in nondiabetic critically ill patients.

**Article Summary:**

Strengths and limitations of this study
- This is the first study to evaluate the association between less tight TIR (70-180 mg/dl) and mortality in critically ill patients.
- We found consistent results in the analyses using logistic regression model and Cox proportional hazard model.
- The primary findings were also observed in the analysis using TIR in the first three days.
- We did not use a continuous glucose monitoring device to calculate TIR.
- Due to the nature of the single-centre study design, the generalizability of the findings might be limited.

## INTRODUCTION

Glycemic control for critically ill patients remains an important issue related to critical care. Although intensive insulin therapy (IIT) had been recommended [1, 2], a large randomized controlled trial (RCT) demonstrated no benefit of IIT for critically ill patients [3]. Therefore, the current recommendation for glycemic control management for critically ill patients has been changed from tight to mild control [4, 5].

Recently, the importance of the percentage of time in targeted blood glucose range (TIR) has been emphasized as a prognostic factor for critically ill patients [6]. In previous studies, TIR 70-140 mg/dL <80% was associated with higher mortality in nondiabetic critically ill patients with HbA1c ≤6.5% [7]. However, current national guidelines recommend the more liberal upper target glucose level of 180 mg/dL [8]. Indeed, the *Surviving Sepsis Campaign Guidelines* suggest the upper limit of the glucose level at ≤180 mg/dL [9]. Furthermore, a systematic review of 36 RCTs and the *American Diabetes Association* also recommend a target glucose range of 140-180 mg/dL for most critically ill patients (grade A) [10]. Despite the clinical consensus on the liberal upper target glucose level of 180 mg/dL, little is known about the association between glycemic control with TIR 70-180 mg/dL and prognosis of critically ill patients. Additionally, it remains unclear whether the optimal TIR for critically ill patients should be maintained at a target upper glucose level of ≤180 mg/dL.

To address the knowledge gap in the literature, we examined the association of TIR 70-180 mg/dL with 28-day mortality among patients admitted to the intensive care unit (ICU). We hypothesized that higher TIR would be associated with a lower risk of 28-day mortality in patients admitted to the ICU.

## METHODS

### Study design and setting

This retrospective observational study used data from the Hitachi General Hospital from 1 January 2016 through 31 October 2019. Hitachi General Hospital, a tertiary care centre in Japan, serves an area with approximately three million residents. Annual emergency department visits are about 27,000 encounters. The Hitachi General Hospital has 18 ICUs and six cardiac care units. Of the 18 ICU beds, eight beds have a 2:1 patient-nurse ratio. The remaining ten beds have a 4:1 patient-nurse ratio. The ICU physicians manage all critically ill patients (both intrinsic and extrinsic) except after cardiovascular surgery. The ICU physicians input clinical information into the ICU database, which automatically registers medical charts as structured data. The study protocol was approved by the Ethics Committee of Hitachi General Hospital (2017-95). The need for informed consent was waived based on the study’s retrospective design.

### Study population

We included all adult ICU admission patients (aged ≥ 20 years). When a patient had multiple admissions, we used the first admission and the first ICU stay during the study period. According to the previous literature, we excluded patients with diabetic ketoacidosis (DKA) or hyperosmolar hyperglycemic syndrome (HHS) [7]. We also excluded patients for whom fewer than nine blood glucose measurements were taken during the ICU stay (i.e., ICU length of stay <3 days) because at least nine blood glucose measurements were necessary to calculate TIR [7]. Moreover, these patients were postoperative patients who left the ICU immediately and did not need glycemic control, or those who died immediately.

### Data collection

We extracted patients’ physical information (age, sex, and body mass index [BMI]), disease severity (acute physiology score [APS] and acute physiology and chronic health evaluation [APACHE] score), disease category, and comorbidity according to the Charlson Comorbidity Index. The APACHE - score was calculated based on vital signs and blood test results measured at ICU admission. Using the measured blood glucose values, the mean blood glucose, glycemic variability, time above range over 220 mg/dL, time below range under 60 or 40 mg/dL, and TIR 70-180 mg/dL were calculated. In addition, we also extracted 28-day mortality, the length of ICU and hospital stay, and patient disposition.

### Glycemic control in the ICU

In the ICU of Hitachi General Hospital, staff members routinely collect blood gas every eight hours (three times per day) from patients with arterial line insertions. All blood glucose was measured using a blood gas analyser (ABL90 FLEX, Radiometer, CPH, DK). Insulin was usually administered as a continuous infusion at a 1 U/mL concentration using a syringe pump with a target range of 70-180 mg/dL. Although the adjustment of the insulin dosage and its timing are made at the intensivist’s discretion, the standard protocol of glycemic control in the ICU is the following: 1) start continuous IV insulin when blood glucose is >180 mg/dL (for patients receiving continuous nutrition); (2) discontinue IV insulin if blood glucose falls <100 mg/dL; and (3) IV dextrose if blood glucose falls ≤70 mg/dL.

### Exposure

The primary exposure was TIR 70-180 mg/dL. The TIR was calculated as the percentage of times the blood glucose met 70-180 mg/dL out of the total number of blood glucose measurements.

### Outcome measures

The primary outcome was 28-day mortality.

### Statistical analysis

Summary statistics were used to describe the characteristics of the study participants. Missing height and weight data occurred in 23.2% of these patients, but there were no missing data or marked outliers in blood glucose data.

According to the previous literature, with stratification by HbA1c ≥6.5mg/dL [7], we examined the association between TIR ≥80% and 28-day mortality using two multivariate models: 1) logistic regression and 2) Cox proportional hazards models. We adjusted for age, sex, APACHE score, Charlson comorbidity index, and the primary diagnosis category (sepsis, cerebrovascular diseases, cardiac diseases, respiratory diseases, gastrointestinal diseases, trauma, postoperative, and others). Kaplan-Meier survival curves were graphed for these populations.

To assess the optimal range of TIR, we repeated the analyses using TIR as a 1) category with 10% incremental of TIR (<60%, 60%-69%, 70%-79%, 80%-89%, and ≥90%) and as a 2) category based on quartiles of patient distribution (<53%, 53%-80%, 81%-93%, and ≥94%; **Supplemental Figure 1**). Furthermore, to assess the linear association of TIR with the outcome, we repeated the analyses using TIR (10% decremental) as a continuous variable. Lastly, because surviving patients were more likely to have stable TIR, we repeated the primary analysis using the TIR in the first three days. A two-sided p-value of <0.05 was considered statistically significant. Statistical analyses were conducted using Stata ver. 15.0 (Stata Corp, TX, USA).

## RESULTS

### Patient characteristics

From 1 January 2016 through 31 October 2019, 2,288 patients were admitted to our ICU. Of these, we excluded 48 patients under the age of 20 years, 43 patients with DKA and HHS, 893 patients with <9 blood glucose measurements, and 74 multiple ICU admissions. The remaining 1,230 patients were eligible for the analysis (**Supplemental Figure 2**). Their mean age was 72 years; 65% were male, with mean BMI of 22.1 (**Table 1**). The mean HbA1c on admission was 6.1%; 250 patients (20.2%) had HbA1c ≥6.5%. No significant differences were found in age, BMI, comorbidity, or, severity (such as APS or APACHE score) between patients with HbA1c <6.5% and patients with HbA1c ≥6.5%. Patients with HbA1c ≥6.5% had significantly higher mean blood glucose levels and glycemic variability than those with HbA1c <6.5%. Mean TIR was 77.7% for patients with HbA1c <6.5% and 49.1% for patients with HbA1c ≥6.5% (p <0.001). Patients with HbA1c ≥6.5% had higher 28-day mortality than those with <6.5% (32.0% vs. 22.7%; p =0.003).

**Table 1.** Patient characteristics, glycemic control metrics, and outcomes, stratified according to HbA1c levels.

|  | All patients<br>(n =1,230) | HbA1c <6.5%<br>(n =980) | HbA1c ≥6.5%<br>(n =250) | P value |
| --- | --- | --- | --- | --- |
| Patient characteristics |  |  |  |  |
| Age, years | 72.0 (±13.9) | 71.8 (±14.3) | 73.1 (±12.3) | 0.19 |
| Male sex | 797 (64.8) | 618 (63.1) | 179 (71.6) | 0.01 |
| Body Mass Index | 22.1 (±3.9) | 22.0 (±4.1) | 22.5 (±3.4) | 0.15 |
| Acute Physiology Score | 13.7 (±6.9) | 13.6 (±6.9) | 14.1 (±7.2) | 0.26 |
| APACHE II score | 18.8 (±7.2) | 18.7 (±7.2) | 19.2 (±7.3) | 0.34 |
| Charlson Comorbidity Index score | 3.8 (±2.1) | 3.8 (±2.2) | 3.6 (±1.9) | 0.1 |
| HbA1c, % | 6.1 (±1.4) | 5.6 (±0.5) | 8.0 (±2.0) | <0.001 |
| Diagnosis |  |  |  | 0.01 |
| Sepsis | 310 (25.2) | 233 (23.8) | 77 (30.8) |  |
| Cerebrovascular diseases | 122 (9.9) | 102 (10.4) | 20 (8.0) |  |
| Cardiac diseases | 96 (7.8) | 71 (7.2) | 25 (10) |  |
| Cardiac arrest | 126 (10.2) | 91 (9.3) | 35 (14) |  |
| Respiratory diseases | 173 (14.1) | 147 (15) | 26 (10.4) |  |
| Gastrointestinal diseases | 58 (4.7) | 47 (4.8) | 11 (4.4) |  |
| Trauma | 63 (5.1) | 49 (5.0) | 14 (5.6) |  |
| Postoperative | 46 (3.7) | 39 (4.0) | 7 (2.8) |  |
| Others | 236 (19.2) | 201 (20.5) | 35 (14) |  |
| Glycemic control metrics |  |  |  |  |
| Mean blood glucose, mg/dL | 163.5 (±40.8) | 154.6 (±32.5) | 198.4 (±50.6) | <0.001 |
| Coefficient of variation, % | 39.3 (±30.7) | 33.7 (±23.2) | 61.3 (±44.0) | <0.001 |
| Time above 220 mg/dL, % | 13.1 (±19.7) | 8.7 (±15.3) | 30.4 (±24.9) | <0.001 |
| Time below 60 mg/dL, % | 0.6 (±2.3) | 0.5 (±2.2) | 0.8 (±2.7) | 0.03 |
| Time below 40 mg/dL, % | 0.2 (±1.3) | 0.2 (±1.2) | 0.4 (±1.7) | 0.03 |
| Time in range 70-180 mg/dL, % | 71.9 (±26.9) | 77.7 (±22.7) | 49.1 (±29.6) | <0.001 |
| Time in range 70-180 mg/dL ≥80% | 644 (52.4) | 586 (59.8) | 58 (23.2) | <0.001 |
| Outcomes |  |  |  |  |
| ICU stay, days | 7.5 (±4.7) | 7.4 (±4.6) | 7.7 (±4.9) | 0.48 |
| Hospital stay, days | 35.2 (±43.2) | 35.7 (±44.2) | 33.5 (±39.0) | 0.49 |
| 28-day mortality | 304 (24.7) | 224 (22.9) | 80 (32.0) | 0.004 |

Data are presented as mean (± standard deviation) or number (percentage). APACHE, Acute Physiology and Chronic Health Evaluation; HbA1c, glycosylated haemoglobin; ICU, Intensive care unit.

### Association of TIR ≥80% with 28-day mortality

Compared to TIR ≥80%, TIR <80% was associated significantly with the worse 28-day mortality in patients with HbA1c <6.5% (unadjusted odds ratio [OR], 2.23 [95%CI 1.65-3.01]; **Table 2** and **Figure 1**). This association remained significant in the multivariate logistic regression model with a similar effect size (adjusted OR, 2.21 [95%CI 1.60-3.05]). By contrast, in patients with HbA1c ≥6.5%, no significant association was found between TIR ≥80% and 28-day mortality in either unadjusted or adjusted logistic regression models. These associations of TIR with 28-day mortality were consistent with results obtained using Cox proportional hazards models (**Supplemental Table 1**).

**Table 2.** Associations between time in range and 28-day mortality with odds ratios and 95% confidence intervals.

| Time in range | HbA1c <6.5% |  |  |  | HbA1c ≥6.5% |  |  |  |
| --- | --- | --- | --- | --- | --- | --- | --- | --- |
|  | Mortality / n | Mortality rate | Unadjusted OR | Adjusted OR† | Mortality / n | Mortality rate | Unadjusted OR | Adjusted OR† |
| Time in range (threshold at 80%) |  |  |  |  |  |  |  |  |
| <80% | 124 / 394 | 32% | 2.23 (1.65-3.01)*** | 2.21 (1.60-3.05)*** | 59 / 192 | 31% | 0.78 (0.42-1.45) | 0.64 (0.30-1.37) |
| ≥80% | 100 / 586 | 17% | 1 (reference) | 1 (reference) | 21 / 58 | 36% | 1 (reference) | 1 (reference) |
| Time in range (10% incremental category) |  |  |  |  |  |  |  |  |
| <60% | 78 / 205 | 38% | 3.76 (2.53-5.58)*** | 3.62 (2.36-5.53)*** | 56 / 160 | 35% | 1.77 (0.71-4.38) | 1.19 (0.40-3.53) |
| 60%-69% | 24 / 86 | 28% | 2.40 (1.39-4.16)** | 2.39 (1.33-4.28)** | 1 / 13 | 8% | 0.27 (0.03-2.49) | 0.13 (0.01-1.42) |
| 70%-79% | 22 / 103 | 21% | 1.67 (0.96-2.88) | 1.63 (0.92-2.91) | 2 / 19 | 11% | 0.39 (0.07-2.10) | 0.14 (0.02-0.97) |
| 80%-89% | 43 / 178 | 24% | 1.96 (1.26-3.06)** | 1.83 (1.14-2.93)* | 14 / 28 | 50% | 3.29 (1.07-10.1)* | 1.81 (0.46-7.06) |
| ≥90% | 57 / 408 | 14% | 1 (reference) | 1 (reference) | 7 / 30 | 23% | 1 (reference) | 1 (reference) |
| Time in range (10% decremental) |  |  | 1.21 (1.14-1.29)*** | 1.20 (1.12-1.28)*** |  |  | 1.02 (0.94-1.12) | 1.08 (0.96-1.21) |
| Time in range (quartile) |  |  |  |  |  |  |  |  |
| Q1 (<53%) | 60 / 156 | 39% | 4.07 (2.55-6.48)*** | 3.73 (2.26-6.14)*** | 53 / 151 | 35% | 1.71 (0.64-4.55) | 1.21 (0.36-4.05) |
| Q2 (53%-80%) | 70 / 260 | 27% | 2.42 (1.57-3.74)*** | 2.28 (1.44-3.61)*** | 6 / 44 | 14% | 0.50 (0.14-1.76) | 0.17 (0.04-0.76)* |
| Q3 (81%-93%) | 55 / 268 | 21% | 1.71 (1.09-2.67)* | 1.57 (0.98-2.53) | 15 / 30 | 50% | 3.17 (0.99-10.1) | 2.06 (0.49-8.75) |
| Q4 (≥94%) | 39 / 296 | 13% | 1 (reference) | 1 (reference) | 6 / 25 | 24% | 1 (reference) | 1 (reference) |

\* P value <0.05; \*\* P value <0.01; \*\*\* P value <0.001.
† Logistic regression adjusted for age, sex, Charlson comorbidity index, APACHE II score, and primary diagnosis category (sepsis, cerebrovascular diseases, cardiac diseases, cardiac arrest, respiratory diseases, gastrointestinal diseases, trauma, postoperative, and others). APACHE, Acute Physiology and Chronic Health Evaluation; HbA1c, glycosylated haemoglobin; OR, odds ratio; CI, confidence interval.

**Figure 1.**
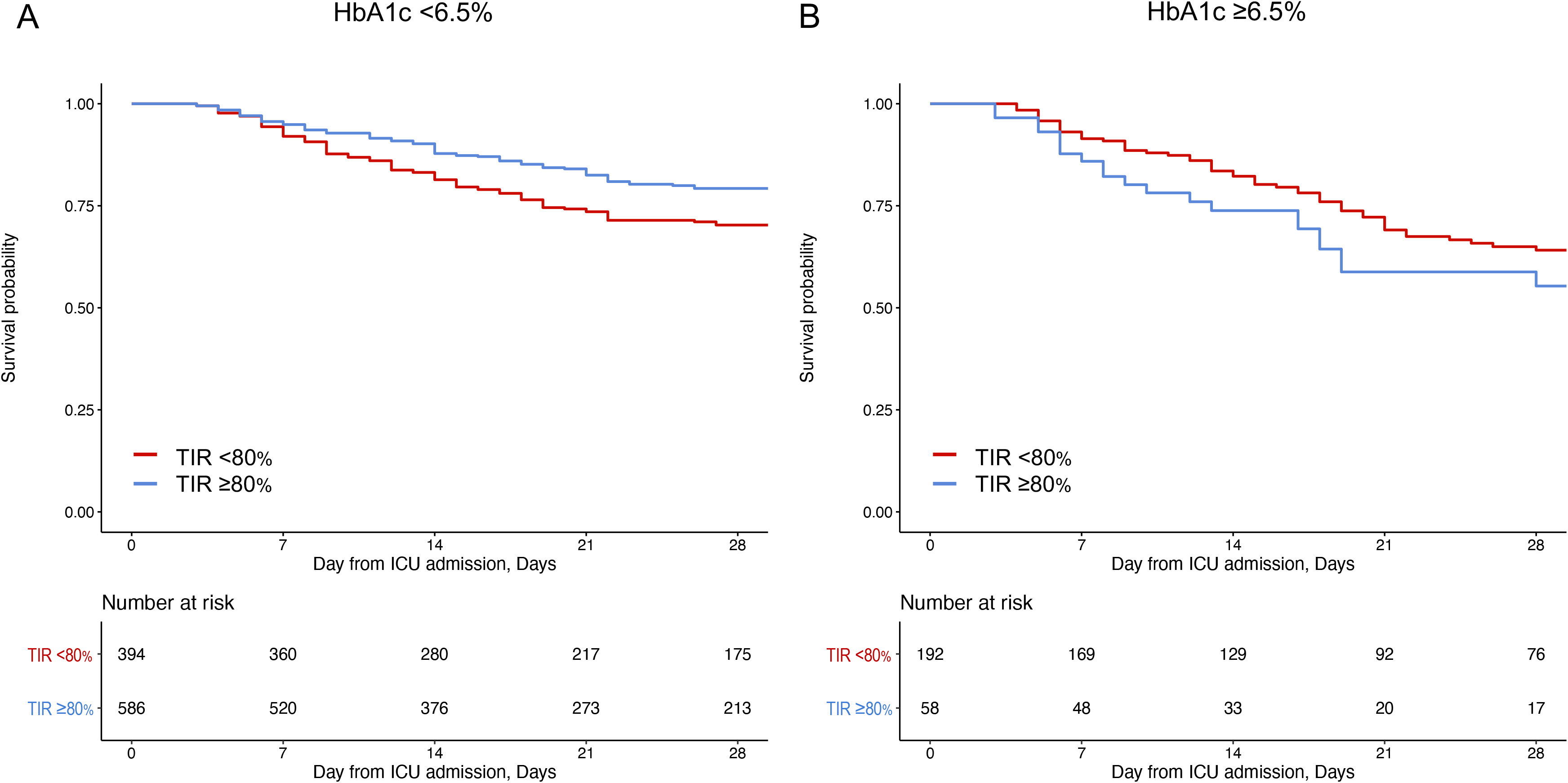
Association of time in range with 28-day survival according to HbA1c. Kaplan-Meier curves for patients with and without TIR <80%; TIR <80% (red) and TIR ≥80% (blue), in patients with HbA1c <6.5% (A), patients with HbA1c ≥6.5% (B). TIR, Time in range; HbA1c, glycosylated haemoglobin.

### Association of TIR category (10% incremental) with 28-day mortality

When using 10% incremental TIR as a categorical variable (<60%, 60%-69%, 70%-79%, 80%-89%, and ≥90%), the lower TIR was associated with the worse 28-day mortality compared to TIR ≥90% in patients with HbA1c <6.5% in both unadjusted and adjusted models (e.g., adjusted OR of TIR <60%, 3.62 [95%CI 2.36-5.53]; **Table 2**). In addition, there was a linear association between 10% TIR decrement and the increased risk of 28-day mortality (adjusted OR per 10% decrement, 1.20 [95%CI 1.12-1.28]). By contrast, no clear association was found between TIR and mortality in patients with HbA1c ≥6.5% in either unadjusted or adjusted logistic regression models. These relations of TIR with 28-day mortality were consistent with results obtained using Cox proportional hazards models (**Supplemental Table 1** and **Supplemental Figure 3**).

### Association of TIR category (quartile category) with 28-day mortality

When using TIR as a categorical variable based on a quartile of patients (<53%, 53%-80%, 81%-93%, and ≥94%), the association between the TIR category and 28-day mortality was similar to those using 10% incremental TIR as a categorical variable. For example, TIR <53% was associated with worse 28-day mortality compared to TIR ≥94% (adjusted OR 3.73 [95%CI 2.26-6.14]; **Table 2** and **Figure 2**).

**Figure 2.**
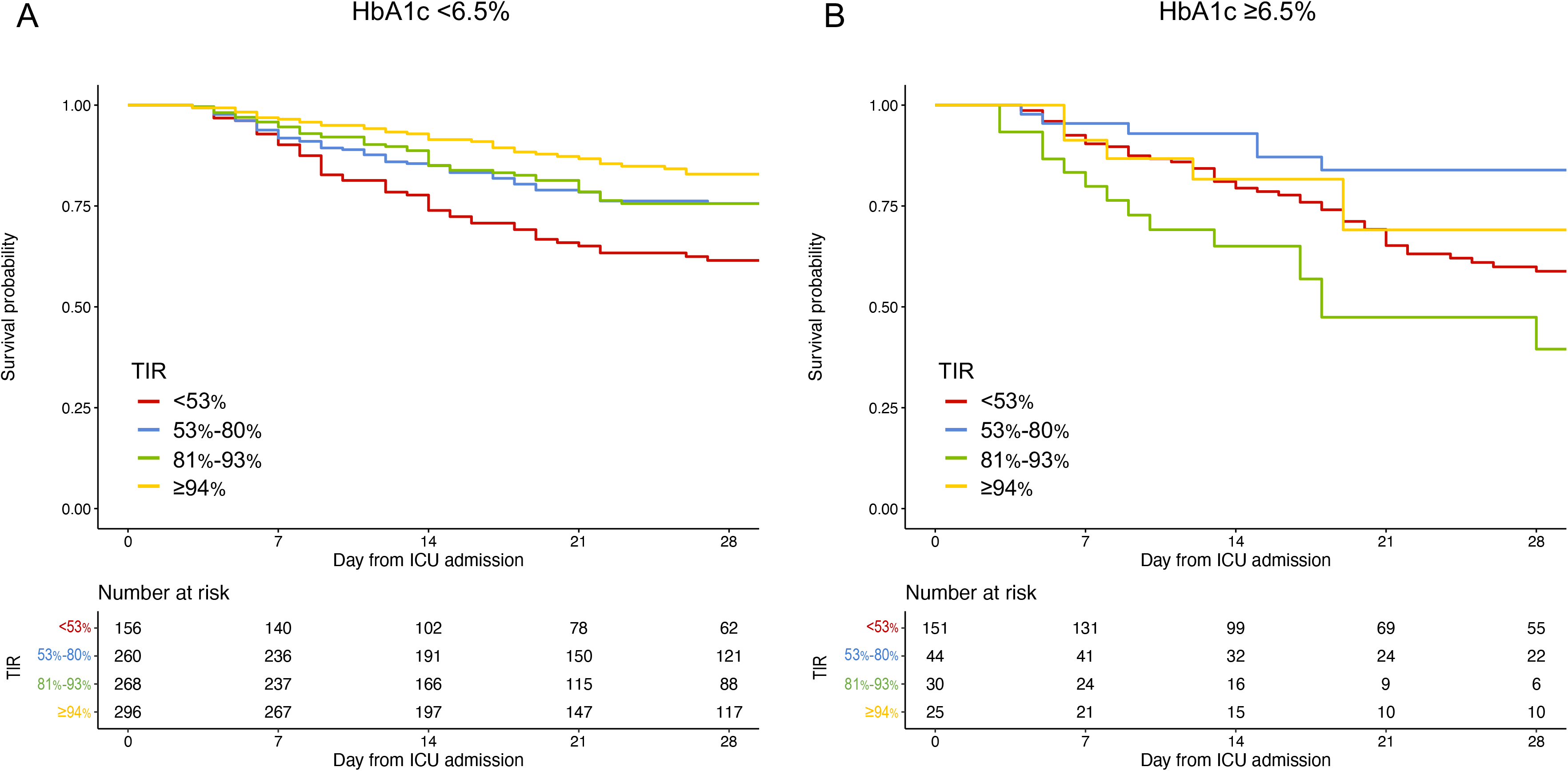
Association of time in range as a categorical variable (quartile) with 28-day survival according to HbA1c. Kaplan-Meier curves for patients with each quartile category of TIR; <53% (red), 53%-80% (blue), 81%-93% (green), and ≥94% (yellow), in patients with HbA1c <6.5% (A), patients with HbA1c ≥6.5% (B). TIR, Time in range; HbA1c, glycosylated haemoglobin.

### Sensitivity analyses using TIR of the first three days

In the analysis using TIR based on the first three days, the overall associations were consistent with those found from the primary analyses. For example, TIR <80% was associated with the worse 28-day mortality in patients with HbA1c <6.5% (e.g., adjusted OR of TIR <80%, 1.88 [95%CI 1.36-2.61]: **Table 3**). Similarly, the lower TIR was associated with the worse 28-day mortality in patients with HbA1c <6.5% (e.g., adjusted OR of TIR <60%, 2.02 [95%CI 1.33-3.06]; **Table 3**).

**Table 3.** Results of sensitivity analyses using time in range of the first three days.

| Time in range | HbA1c <6.5% |  |  |  | HbA1c ≥6.5% |  |  |  |
| --- | --- | --- | --- | --- | --- | --- | --- | --- |
|  | Mortality / n | Mortality rate | Unadjusted OR | Adjusted OR† | Mortality / n | Mortality rate | Unadjusted OR | Adjusted OR† |
| Time in range (threshold at 80%) |  |  |  |  |  |  |  |  |
| <80% | 144 / 491 | 29% | 2.12 (1.56-2.89)*** | 1.88 (1.36-2.61)*** | 67 / 210 | 32% | 0.97 (0.47-2.00) | 0.56 (0.23-1.37) |
| ≥80% | 80 / 489 | 16% | 1 (reference) | 1 (reference) | 13 / 40 | 33% | 1 (reference) | 1 (reference) |
| Time in range (10% incremental category) |  |  |  |  |  |  |  |  |
| <60% | 84 / 261 | 32% | 2.32 (1.57-3.42)*** | 2.02 (1.33-3.06)*** | 56 / 171 | 33% | 1.32 (0.52-3.33) | 0.65 (0.21-1.97) |
| 60%-69% | 20 / 92 | 22% | 1.36 (0.77-2.41) | 1.16 (0.63-2.14) | 4 / 15 | 27% | 0.99 (0.23-4.15) | 0.25 (0.05-1.37) |
| 70%-79% | 40 / 138 | 29% | 2.00 (1.25-3.19)** | 1.90 (1.15-3.13)* | 7 / 24 | 29% | 1.12 (0.32-3.84) | 0.37 (0.08-1.68) |
| 80%-89% | 25 / 165 | 15% | 0.87 (0.52-1.46) | 0.88 (0.51-1.51) | 6 / 14 | 43% | 2.04 (0.52-8.00) | 1.09 (0.20-6.07) |
| ≥90% | 55 / 324 | 17% | 1 (reference) | 1 (reference) | 7 / 26 | 27% | 1 (reference) | 1 (reference) |
| Time in range (10% decremental) |  |  | 1.11 (1.05-1.16)*** | 1.09 (1.03-1.15)** |  |  | 1.01 (0.93-1.10) | 1.01 (0.91-1.12) |
| Time in range (quartile) |  |  |  |  |  |  |  |  |
| Q1 (<53%) | 61 / 187 | 33% | 2.28 (1.42-3.65)*** | 2.07 (1.25-3.43)** | 50 / 153 | 33% | 1.34 (0.54-3.66) | 0.66 (0.21-2.05) |
| Q2 (53%-80%) | 83 / 304 | 27% | 1.91 (1.27-2.89)*** | 1.61 (1.04-2.50)* | 17 / 57 | 30% | 1.19 (0.44-3.50) | 0.52 (0.15-1.76) |
| Q3 (81%-93%) | 25 / 165 | 15% | 1.34 (0.90-1.99) | 1.52 (0.86-2.01) | 6 / 14 | 43% | 1.41 (0.48-4.39) | 0.56 (0.15-2.10) |
| Q4 (≥94%) | 55 / 324 | 17% | 1 (reference) | 1 (reference) | 7 / 26 | 27% | 1 (reference) | 1 (reference) |

\* P value <0.05; \*\* P value <0.01; \*\*\* P value <0.001.
† Logistic regression adjusted for age, sex, Charlson comorbidity index, APACHE II score, and primary diagnosis category (sepsis, cerebrovascular diseases, cardiac diseases, cardiac arrest, respiratory diseases, gastrointestinal diseases, trauma, postoperative, and others). APACHE, Acute Physiology and Chronic Health Evaluation; HbA1c, glycosylated haemoglobin; OR, odds ratio.

## DISCUSSION

This study of 1,230 ICU patients found that TIR 70-180 mg/dL was associated significantly with 28-day mortality in critically ill patients with HbA1c <6.5%, although no apparent association was found in those with HbA1c ≥6.5%. While the causal relationship cannot be determined, based on the analyses conducted using several cut-off values, the optimal range of TIR may be >90%, or >80% is also reasonable for patients with HbA1c <6.5%. Furthermore, similar findings obtained using TIR based on the first three days support the use of TIR as a prognostic marker for critically ill patients.

The observed relation of higher TIR with lower mortality in critically ill patients was consistent with those of earlier studies [7, 11]. Krinsley et al. examined mortality between the high-TIR and low-TIR groups and found significantly lower mortality in the high-TIR group than in the low-TIR group among non-diabetic patients (8.5% vs. 15.8%, p <0.001), although no difference was found in mortality between groups among diabetic patients (16.1% vs. 14.4%, p =0.60) [11]. Similarly, Lanspa et al. found that the relation between TIR >80% and mortality was likely to depend on HbA1c rather than on whether the patient had diabetes mellitus [7].

Earlier studies of critically ill patients have described an association between TIR and mortality with a target range of 70-140 mg/dL for blood glucose [7, 11]. Whereas these earlier studies provided concrete evidence on the glucose management for ICU patients, the current national guidelines recommend the more liberal upper target glucose level of 180 mg/dL [10]. Moreover, few studies have addressed survival bias, by which surviving patients were more likely to have stable TIR [12]. In this context, our findings based on the target range of the glucose level of 70-180 mg/dL are expected to extend these earlier studies, by demonstrating the optimal TIR and the TIR-outcome relation using data for the first three days. Moreover, these findings facilitate future trials aimed at determining the optimal TIR range for ICU patients.

Critically ill patients can easily become hyperglycemic because of various factors [13, 14]. Glucose is a pro-inflammatory mediator with mechanisms such as increasing plasma interleukin-8 levels [15], increasing nuclear factor kappa-light-chain-enhancer of activated B cells [16], and increasing plasma matrix metalloproteinase (MMP) −2 and MMP-9 levels [17]. Although hyperglycemia rarely causes death directly, prolonged hyperglycemia can contribute to morbid conditions such as sepsis and post-intensive care syndrome by causing a toxic cellular environment and impaired immune function [18]. By contrast, hypoglycemia is an iatrogenic complication that should be avoided. The NICE-SUGAR trial, which defined hypoglycemia, reported an increased risk of death in patients with moderate to severe hypoglycemia compared to those without [3]. Hypoglycemia presents an independent risk of death, as well as risks of convulsions and arrhythmias [19, 20]. The management of glycemic control to avoid hyperglycemia and hypoglycemia, as clinicians have attempted for many decades, is nothing less than maintaining high TIR management. Therefore, the finding that high TIR was associated significantly with low mortality was clinically plausible.

Importantly, the association between TIR 70-180 mg/dL and mortality was found only in patients with HbA1c <6.5% on admission: not in those with ≥6.5%. Several studies have revealed similar associations [7, 11]. Indeed, unlike patients with diabetes mellitus, TIR 70-140 mg/dL >80% was not associated with mortality in critically ill patients without diabetes mellitus [11]. Possible explanations include that patients with HbA1c ≥6.5% on admission were more likely to have been chronically hyperglycemic. Chronic hyperglycemia may attenuate the extent of cellular toxicity in acute hyperglycemia by downregulating the glucose transporter (GLUT) 1 and GLUT 3 transporters [21]. In addition, patients with a high HbA1c on admission may be less likely to be harmed by a low TIR because they are relatively less harmed by exposure to hyperglycemia [22].

### Potential Limitations

Several potential limitations exist for this study. First, unmeasured confounding factors related to assessment of the association of TIR with outcomes may exist because this was a retrospective observational study. Second, this study calculated TIR using glucose measured using a blood gas analyser every eight hours. This may not give an accurate representation of actual TIR when measured, for example, with continuous glucose monitoring. Third, because this study was conducted in a tertiary care hospital in Japan, the generalizability of the findings may be limited. Yet, these findings were consistent with previous studies [7,11] and the observed association may not be substantially varied across hospitals.

## CONCLUSIONS

Our study demonstrated that lower TIR 70-180 mg/dL was associated with higher mortality in critically ill patients with good antecedent glycemic control. Further validation studies are needed, but for glycemic control at a range of 70-180 mg/dL, TIR ≥90%, at least ≥80%, may be the target for critically ill patients with HbA1c <6.5%.

## Data Availability

The datasets used and analysed during the current study are available from the corresponding author on reasonable request.

**List of abbreviations**
APACHE: Acute physiology and chronic health evaluation
APS: Acute physiology score
BMI: Body mass index
CI: Confidence interval
DKA: Diabetic ketoacidosis
GLUT: Glucose transporter
HbA1c: Glycosylated haemoglobin
HHS: Hyperosmolar hyperglycemic syndrome
HR: Hazard ratio
IUC: Intensive care unit
IIT: Intensive insulin therapy
MMP: Matrix metalloproteinase
OR: Odds ratio
RCT: Randomized controlled trial
TIR: Time in range

## Declarations

### Ethics approval and consent to participate

This retrospective observational study was approved by the Ethics Committee of Hitachi General Hospital (2017-95). The need for informed consent was waived based on the study’s retrospective design.

### Consent for publication

Not applicable.

### Authors’ contributions

HN, TS, and TG developed the analysis plan. HN undertook the main analysis and composed the first draft of the paper with supervision from TG. All other authors made important revisions. All authors had full access to all the data used for the study. All take responsibility for the data integrity and the data analysis accuracy.

## Acknowledgements

Not applicable.

## Additional files

**Supplemental Table 1**

***File name and format:*** TIR_Table_s 1. docx

***Title:*** Unadjusted and adjusted associations between time in range and 28-day mortality using COX proportional hazard model

***Description:***

* P value <0.05; ** P value <0.01; *** P value <0.001.

† Cox proportional hazards model adjusted for age, sex, Charlson comorbidity index, APACHE II score, and primary diagnosis category (sepsis, cerebrovascular diseases, cardiac diseases, cardiac arrest, respiratory diseases, gastrointestinal diseases, trauma, postoperative, and others). APACHE, Acute Physiology and Chronic Health Evaluation; HbA1c, glycosylated haemoglobin; HR, hazard ratio.

**Supplemental Figure 1**

***File name and format:*** TIR_Figure_s1.pdf

***Title:*** Distribution of time in range

***Description:*** The quartiles of TIR 70-180 mg/dL were as follow, lower quartile, 53.2%; median, 80.7%; upper quartile, 93.3%. TIR, Time in range.

**Supplemental Figure 2**

***File name and format:*** TIR Figure_s2.pdf

***Title:*** Study flow

**Supplemental Figure 3**

***File name and format:*** TIR_Figure_s3.pdf

***Title:*** Association of time in range as 10% incremental category with 28-day mortality according to HbA1c

***Description:*** Kaplan-Meier curves for patients with each 10% incremental category of TIR; <60% (red), 60%-69% (blue), 70%-79% (green), 80%-89% (yellow), and ≥90% (gray), in patients with HbA1c <6.5% (A), patients with HbA1c ≥6.5% (B). TIR, Time in range; HbA1c, glycosylated haemoglobin.

## REFERENCES

1. Van den Berghe G, Wouters P, Weekers F, Verwaest C, Bruyninckx F, Schetz M, Vlasselaers D, Ferdinande P, Lauwers P, Bouillon R: Intensive insulin therapy in critically ill patients. N Engl J Med 2001, 345(19):1359–1367.

2. Van den Berghe G, Wilmer A, Hermans G, Meersseman W, Wouters PJ, Milants I, Van Wijngaerden E, Bobbaers H, Bouillon R: Intensive insulin therapy in the medical ICU. N Engl J Med 2006, 354(5):449–461.

3. Finfer S, Chittock DR, Su SY, Blair D, Foster D, Dhingra V, Bellomo R, Cook D, Dodek P, Henderson WR et al: Intensive versus conventional glucose control in critically ill patients. N Engl J Med 2009, 360(13):1283–1297.

4. American Diabetes Association: Standards of medical care in diabetes--2014. Diabetes Care 2014, 37 Suppl 1:S14-80.

5. Qaseem A, Chou R, Humphrey LL, Shekelle P: Inpatient glycemic control: best practice advice from the Clinical Guidelines Committee of the American College of Physicians. Am J Med Qual 2014, 29(2):95–98.

6. Aramendi I, Burghi G, Manzanares W: Dysglycemia in the critically ill patient: current evidence and future perspectives. Rev Bras Ter Intensiva 2017, 29(3):364–372.

7. Lanspa MJ, Krinsley JS, Hersh AM, Wilson EL, Holmen JR, Orme JF, Morris AH, Hirshberg EL: Percentage of Time in Range 70 to 139 mg/dL Is Associated With Reduced Mortality Among Critically Ill Patients Receiving IV Insulin Infusion. Chest 2019, 156(5):878–886.

8. Jacobi J, Bircher N, Krinsley J, Agus M, Braithwaite SS, Deutschman C, Freire AX, Geehan D, Kohl B, Nasraway SA et al: Guidelines for the use of an insulin infusion for the management of hyperglycemia in critically ill patients. Crit Care Med 2012, 40(12):3251–3276.

9. Rhodes A, Evans LE, Alhazzani W, Levy MM, Antonelli M, Ferrer R, Kumar A, Sevransky JE, Sprung CL, Nunnally ME et al: Surviving Sepsis Campaign: International Guidelines for Management of Sepsis and Septic Shock: 2016. Intensive Care Med 2017, 43(3):304–377.

10. American Diabetes Association: 13. Diabetes Care in the Hospital. Diabetes Care 2016, 39 Suppl 1:S99-104.

11. Krinsley JS, Preiser JC: Time in blood glucose range 70 to 140 mg/dl >80% is strongly associated with increased survival in non-diabetic critically ill adults. Crit Care 2015, 19:179.

12. Sechterberger MK, Luijf YM, Devries JH: Poor agreement of computerized calculators for mean amplitude of glycemic excursions. Diabetes Technol Ther 2014, 16(2):72–75.

13. McCowen KC, Malhotra A, Bistrian BR: Stress-induced hyperglycemia. Crit Care Clin 2001, 17(1):107–124.

14. Dungan KM, Braithwaite SS, Preiser JC: Stress hyperglycemia. Lancet 2009, 373(9677):1798–1807.

15. Chettab K, Zibara K, Belaiba SR, McGregor JL: Acute hyperglycemia induces changes in the transcription levels of 4 major genes in human endothelial cells: macroarrays-based expression analysis. Thromb Haemost 2002, 87(1): 141-148.

16. Yorek MA, Dunlap JA: Effect of increased concentration of D-glucose or L-fucose on monocyte adhesion to endothelial cell monolayers and activation of nuclear factor-kappaB. Metabolism 2002, 51(2):225–234.

17. Aljada A, Ghanim H, Mohanty P, Syed T, Bandyopadhyay A, Dandona P: Glucose intake induces an increase in activator protein 1 and early growth response 1 binding activities, in the expression of tissue factor and matrix metalloproteinase in mononuclear cells, and in plasma tissue factor and matrix metalloproteinase concentrations. Am J Clin Nutr 2004, 80(1):51–57.

18. Stevens RD, Dowdy DW, Michaels RK, Mendez-Tellez PA, Pronovost PJ, Needham DM: Neuromuscular dysfunction acquired in critical illness: a systematic review. Intensive Care Med 2007, 33(11):1876–1891.

19. Preiser JC, Devos P, Ruiz-Santana S, Melot C, Annane D, Groeneveld J, Iapichino G, Leverve X, Nitenberg G, Singer P et al: A prospective randomised multi-centre controlled trial on tight glucose control by intensive insulin therapy in adult intensive care units: the Glucontrol study. Intensive Care Med 2009, 35(10):1738–1748.

20. Hermanides J, Bosman RJ, Vriesendorp TM, Dotsch R, Rosendaal FR, Zandstra DF, Hoekstra JB, DeVries JH: Hypoglycemia is associated with intensive care unit mortality. Crit Care Med 2010, 38(6):1430–1434.

21. Klip A, Tsakiridis T, Marette A, Ortiz PA: Regulation of expression of glucose transporters by glucose: a review of studies in vivo and in cell cultures. Faseb j 1994, 8(1):43–53.

22. Krinsley JS, Fisher M: The diabetes paradox: diabetes is not independently associated with mortality in critically ill patients. Hosp Pract 2012, 40(2):31–35.

